# Rare Variant Aggregation in 148,508 Exomes Identifies Genes Associated with Proxy Alzheimer’s disease/Dementia

**DOI:** 10.1101/2021.10.17.21265070

**Authors:** Douglas P Wightman, Jeanne E Savage, Christiaan A de Leeuw, Iris E Jansen, Danielle Posthuma

## Abstract

Proxy phenotypes allow for the utilization of genetic data from large population cohorts to analyze late-onset diseases by using parental diagnoses as a proxy for genetic disease risk. Proxy phenotypes based on parental diagnosis status have been used in previous studies to identify common variants associated with Alzheimer’s disease. As of yet, proxy phenotypes have not been used to identify genes associated with Alzheimer’s disease through rare variants. Here we show that a proxy Alzheimer’s disease/dementia phenotype can capture known Alzheimer’s disease risk genes through rare variant aggregation. We generated a proxy Alzheimer’s disease/dementia phenotype for 148,508 unrelated individuals of European ancestry in the UK biobank in order to perform exome-wide rare variant aggregation analyses to identify genes associated with proxy Alzheimer’s disease/dementia. We identified four genes significantly associated with the proxy phenotype, three of which were significantly associated with proxy Alzheimer’s disease/dementia in an independent replication cohort consisting of 197,506 unrelated individuals of European ancestry in the UK biobank. All three of the replicated genes have been previously associated with clinically diagnosed Alzheimer’s disease (*SORL1, TREM2*, and *TOMM40/APOE*). We show that proxy Alzheimer’s disease/dementia can be used to identify genes associated with Alzheimer’s disease through rare variant aggregation.

## Introduction

Rare variants (minor allele frequency (MAF) <0.01) contributing to Alzheimer’s disease (AD) have frequently been identified, first by family-based linkage studies^1–3^, and later by exome sequencing^4,5^ and whole gene sequencing^6,7^. Through these methods multiple genes have been reliably associated with AD through rare variants^8,9^. The sample sizes for these studies generally range from a few thousand individuals^7^ to tens of thousands^10^. Studies with larger sample sizes are more likely to observe rarer variants which provides greater power to conduct rare-variant analyses. Very rare (MAF<1×10^−4^) variants are of particular interest because they are more likely to have a larger impact on the protein of interest^11^, as deleterious variants are likely to undergo negative selection. Due to the relatively late-onset of AD, very few patients are included in large biobank cohorts so large clinically diagnosed AD cohorts have to be generated through patient recruitment. This process is time-consuming and financially costly. Estimation of a proxy AD phenotype may allow for the utilisation of large biobank cohorts to identify variants and genes associated with AD through rare variants. The first description of a proxy AD phenotype based on familial AD status was described in Liu *et al*. (2017)^12^ and later a common variant driven genome-wide association meta-analysis was performed by Marioni *et al*. (2018)^13^, which included a proxy AD phenotype for the UK biobank (UKB) participants. Both of these studies used a case-control design for the proxy phenotype.

Jansen *et al*. (2019)^14^ generated a quantitative proxy phenotype for UKB participants in order to include them with clinically defined cases and controls in a genome-wide meta-analysis of common variation in AD. This quantitative proxy AD/dementia phenotype was based on whether the genotyped individual was diagnosed with any form AD and how many of their parents have an “Alzheimer’s disease/dementia” diagnosis. The contribution of the parental AD/dementia diagnosis was weighed by the ages of the parents. Proxy phenotypes are more diluted phenotypes compared to phenotypes based on clinical diagnosis because genetic risk variants can be lost when alleles are transmitted from parent to offspring. Jansen and colleagues^14^ showed that the power lost due to the diluted phenotype was compensated for by the large sample size of 376,113 individuals. The genetic correlation between the proxy AD/dementia and clinically diagnosed AD was high (r_g_□ = □0.81), which showed that the proxy phenotype was able to capture a large amount of the genetic signal of AD. In the current study, we aimed to apply this same proxy phenotype to rare variant analyses to determine if the proxy phenotype can recapture AD associated variants and genes using rare-variants. In the process, we aim to identify additional genes and variants which may be of interest to AD.

## Results

### Variant aggregation analysis

We performed genome-wide gene-level variant aggregation analyses for 4 variant categories in 148,508 unrelated individuals of European ancestry with a proxy score for AD/dementia. The proxy phenotype was created based on the method described in Jansen *et al*. (2019)^14^, where individuals were assigned a score from 0-2 based on their own AD diagnosis, their reported parental AD/dementia status, and their parents’ age (see **Methods**). Of the 148,508 individuals, 22,080 individuals had at least one parent with AD/dementia or an AD diagnosis themselves. We annotated the rare variants (MAF<0.01) present in these individuals and grouped the variants based on the predicted impact of the variants. The most impactful variants were the high confidence predicted loss-of-function (HiC pLOF) and the least impactful variants were the missense variants. The two intermediate groups were the predicted loss-of-function (pLOF) variants and the high confidence missense variants defined by a REVEL score >50 (REVEL>50). The four variant groups were combined into 4 variant categories: HiC pLOF variants alone (HiC pLOF), all pLOF variants (pLOF), all pLOF variants plus high confidence missense variants (pLOF+REVEL>50), and all pLOF variants plus all missense variants (pLOF+missense). We then performed variant aggregation analyses using the variants present in those 4 categories using SKAT-O. Batch, sex, age, and the first 10 ancestry principal components were used as covariates. Variant aggregation analyses using different variant categories were chosen to limit the analyses to variants with high predicted impacts on protein function to aid interpretation of significant associations. Variants overlapped across the categories to maximize the number of variants in the analyses.

The number of variants in each variant category grows as less impactful variants are included, this also increases the number of tested genes where at least one variant maps to the gene (**Table 1**). The power to observe associations increased with increased number of variants, and this was reflected in the increasing genomic inflation factors as each variant category increases the number of tested genes and variants **(Supplementary Table 1)**. The genomic inflation factors of the variant aggregation analyses were less than one in all four variant categories (HiC pLOF=0.9079775, pLOF=0.9224255, pLOF+REVEL>50=0.9278713, pLOF+missense=0.9443691). The low genomic inflation factors suggest that the signal within the data is sparse, especially in the pLOF variant categories where numbers of variants within a gene were low. Despite the low genomic inflation, four unique genes were found to be significantly associated with proxy AD/dementia (**Figure 1**). As a comparison, we repeated the same variant aggregation analysis, except only using synonymous variants identified by VEP, which resulted in no significantly associated genes after Bonferroni correction and a genomic inflation factor of 0.91. The genomic inflation factor of the synonymous variant analysis was lower than all of the other analyses except the HiC pLOF analysis, this suggests that using variants with clearer functional impacts leads to more association signal in the aggregation analyses. We also performed gene-set analysis by aggregating the variants in genes included in the MSigDB v7.0 gene-sets. Initially, we identified 33 gene-sets which were significantly associated with the proxy AD/dementia phenotype after Bonferroni correction. However, only two gene-sets were even nominally associated after the removal of the larger *APOE* region (GRCh38: 19:40,000,000-50,000,000) and the four significant genes (**Supplementary Note**). Both of the nominally associated gene-sets after removal of the *APOE* region and the four significant genes had a P-value of 0.03. This also suggests that the association signal outside of the four significant genes was sparse.

**Table 1:** The number of variants and genes included in each variant aggregation analysis. The numbers in brackets represent the potential number of genes and variants in the analysis if genes were not excluded due to cumulative allele frequency filtering (<0.0001).

|  | HiC pLOF | pLOF | pLOF + REVEL<br>>50 | pLOF+<br>missense |
| --- | --- | --- | --- | --- |
| # GENES | 6749 (15572) | 11037 (18071) | 16010 (18508) | 18425 (18605) |
| # VARIANTS | 131192<br>(179501) | 246005<br>(295073) | 849567<br>(869322) | 3711893<br>(3712943) |

**Figure 1:**
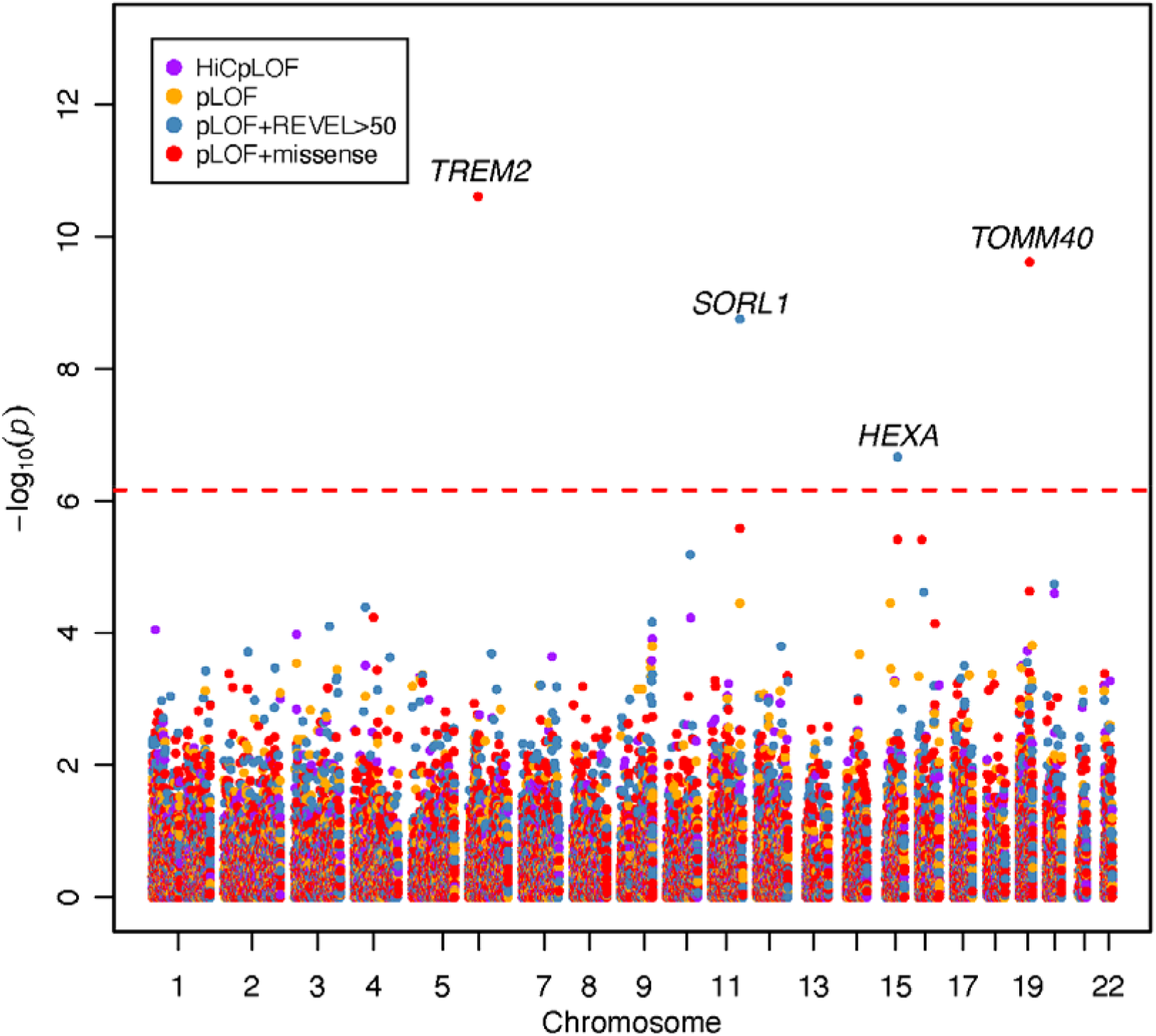
Manhattan plot of the four variant aggregation analyses highlights four significantly associated genes (*TREM2, SORL1, HEXA*, and *TOMM40*). Each point represents a gene in one of the four variant aggregation analyses (HiCpLOF, pLOF, pLOF+REVEL>50, pLOF+missense). The dashed line represents the threshold of significance after correction for all genes tested across all four variant categories.

### Significant Genes, Impact of Singletons, and Replication

*TREM2, TOMM40, SORL1*, and *HEXA* were the four unique genes which reached significance in the SKAT-O^15^ variant aggregation analyses after Bonferroni correction for the number of genes and variant categories (*P*<6.78×10^−7^) **(Table 2)**. *TOMM40, TREM2*, and *SORL1* have been identified in previous rare variant and common variant genome-wide association studies of AD^8,14,16^. *HEXA* has not been previously identified in any rare variant or common variant genome-wide association studies of AD. No significantly associated genes were identified with either HiC pLOF or all pLOF variants alone. *SORL1* and *HEXA* were significantly associated with proxy AD/dementia when testing all pLOF and high confidence missense variants (pLOF+REVEL >50). *TREM2* and *TOMM40* were significantly associated when testing all pLOF and missense variants (pLOF+missense). *TOMM40* is located in relatively proximity to *APOE* so we repeated the analyses of the four associated genes while adding *APOE* ε4 status as an additional covariate. Only the association of *TOMM40* was affected by the additional covariate (*P*=0.043) **(Supplementary Table 2)**, which suggests that the association of *TOMM40* is mediated through *APOE*. We also identified genes from the variant aggregation analyses at Benjamini–Hochberg false-discovery rate (BH-FDR) of 10% in each of the 4 analyses (**Supplementary Note**).

**Table 2:**
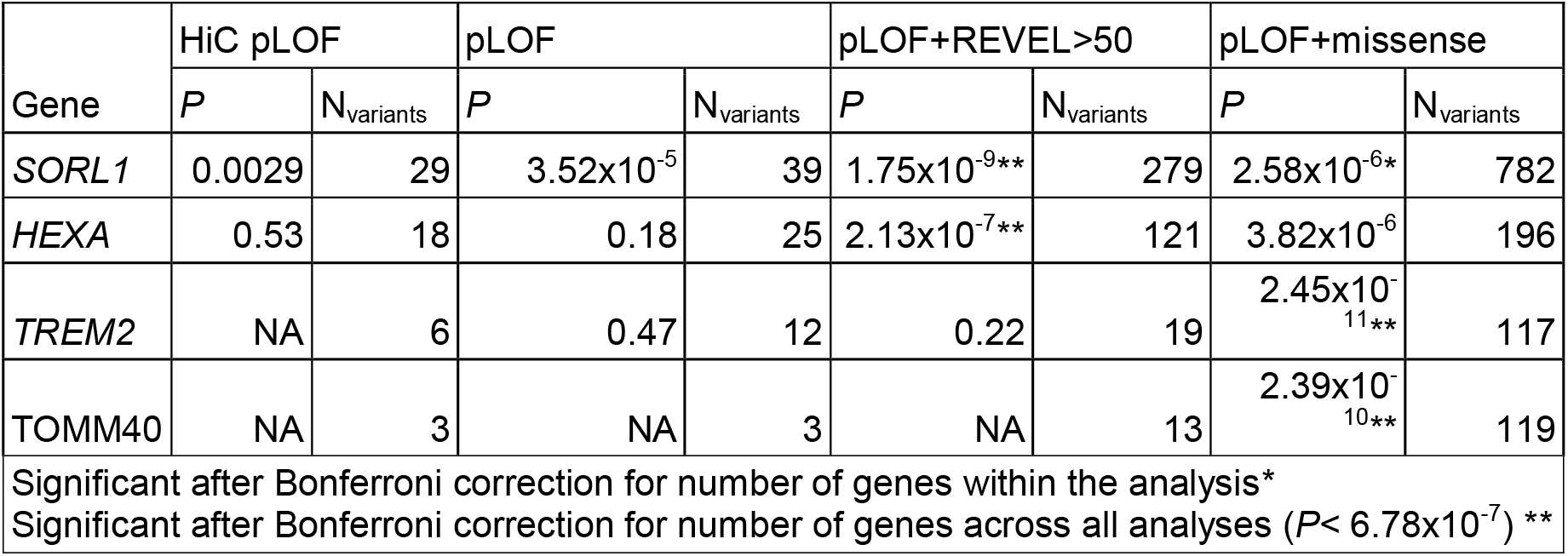
The results from the variant aggregation analyses (SKAT-O) of the four genes (*SORL1, HEXA, TREM2*, and *TOMM40*) which reached significance across the four variant categories (HiC pLOF, pLOF. pLOF+REVEL>50, and pLOF+missense).

| Gene | HiC pLOF |  | pLOF |  | pLOF+REVEL>50 |  | pLOF+missense |  |
| --- | --- | --- | --- | --- | --- | --- | --- | --- |
|  | *P* | *N*<sub>variants</sub> | *P* | *N*<sub>variants</sub> | *P* | *N*<sub>variants</sub> | *P* | *N*<sub>variants</sub> |
| *SORL1* | 0.0029 | 29 | $3.52 \times 10^{-5}$ | 39 | $1.75 \times 10^{-9**}$ | 279 | $2.58 \times 10^{-6*}$ | 782 |
| *HEXA* | 0.53 | 18 | 0.18 | 25 | $2.13 \times 10^{-7**}$ | 121 | $3.82 \times 10^{-6}$ | 196 |
| *TREM2* | NA | 6 | 0.47 | 12 | 0.22 | 19 | $2.45 \times 10^{-11**}$ | 117 |
| *TOMM40* | NA | 3 | NA | 3 | NA | 13 | $2.39 \times 10^{-10**}$ | 119 |

Significant after Bonferroni correction for number of genes within the analysis\*
Significant after Bonferroni correction for number of genes across all analyses ( $P < 6.78 \times 10^{-7}$ ) \*\*

To investigate the impact of moderately associated variants (*P*<1×10^−4^), singletons, and low minor allele count variants (MAC) on the association of the four significant genes, three additional variant aggregation analyses were performed for each of the four significant genes. The three additional analyses were the same as previously described except the moderately associated variants removed, all singletons removed, and all variants with MAC<5 removed (**Supplementary Table 3**). Removing all singletons from the analyses did not cause any gene to lose significance, with *SORL1* being the only gene affected by the loss of singletons (P-value increase from 1.95×10^−9^ to 1.29×10^−7^). However, removing the moderately associated variants caused all genes to lose significance. Removal of all variants with MAC<5 only caused *SORL1* to lose significance (P-value increase from 1.95×10^−9^ to 1.45×10^−5^) suggesting that only *SORL1* is sensitive to the exclusion of very rare variants. This analysis shows that the associations of *TREM2, HEXA*, and *TOMM40* with proxy AD/dementia was driven by variants with moderate associations (*P*<1×10^−4^) and MAC>5, where *SORL1* was influenced by low MAC variants (<5) and moderately associated variants, but not singletons.

We repeated the variant aggregation analyses for *TREM2, SORL1, TOMM40*, and *HEXA* using exome sequencing data of the remaining 197,506 unrelated individuals of European ancestry in the UKB not included in the discovery analysis. *TREM2, SORL1*, and *TOMM40* reached significance in the replication dataset after Bonferroni correction for the number of genes and analyses (*P*<6.78×10^−7^); however, *HEXA* did not reach nominal significance in either the pLOF+REVEL>50 or pLOF+missense analyses (**Table 3**). This suggests that HEXA is unlikely to be associated with proxy AD/dementia. Further discussion of the *HEXA* variant aggregation analyses in the discovery and replication datasets is available in the **Supplementary Note**.

**Table 3:**
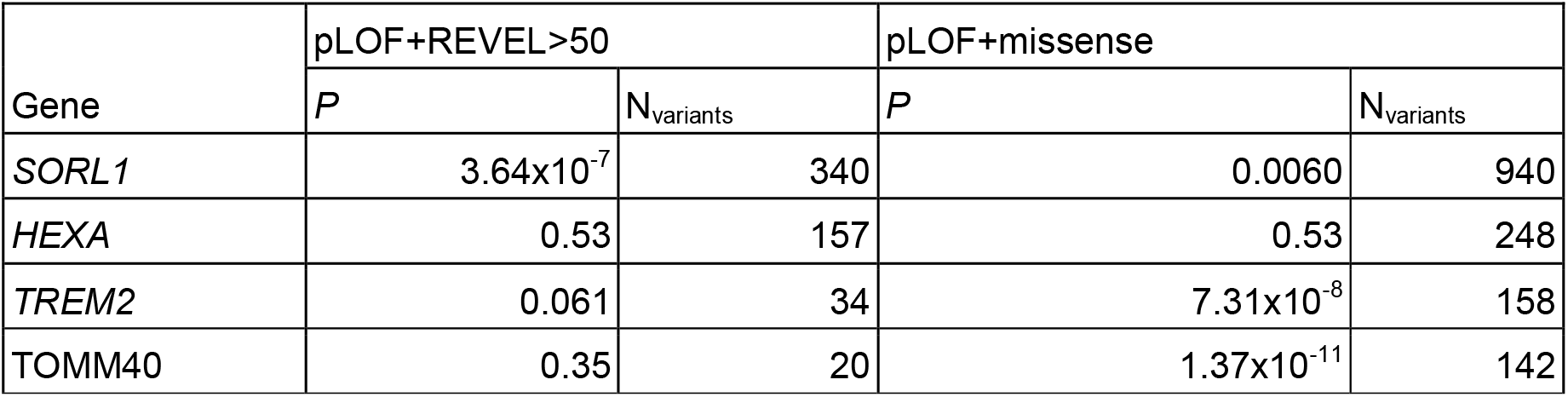
The results from the replication variant aggregation analyses (SKAT-O) of the four genes (*SORL1, HEXA, TREM2*, and *TOMM40*) which reached significance in the discovery variant aggregation analyses.

| Gene | pLOF+REVEL>50 |  | pLOF+missense |  |
| --- | --- | --- | --- | --- |
|  | *P* | *N*<sub>variants</sub> | *P* | *N*<sub>variants</sub> |
| *SORL1* | $3.64 \times 10^{-7}$ | 340 | 0.0060 | 940 |
| *HEXA* | 0.53 | 157 | 0.53 | 248 |
| *TREM2* | 0.061 | 34 | $7.31 \times 10^{-8}$ | 158 |
| *TOMM40* | 0.35 | 20 | $1.37 \times 10^{-11}$ | 142 |

### Variants of Interest

In order to highlight variants of interest within *TREM2* and *SORL1*, we investigated all variants in these genes which were moderately associated (*P*<1×10^−4^) with proxy AD/dementia in the discovery and replication analyses. Discussion of the moderately associated variants within *HEXA* and *TOMM40* is available in the **Supplementary Note**. Due to the limited effect of the singletons on the variant aggregation analyses, we have restricted discussion of moderately associated variants to variants with a MAC>1.

*TREM2* was significantly associated with the phenotype in the pLOF+missense variant category in the discovery dataset (*P*=2.45×10^−11^, N_variants_=117) and replication dataset (*P*=7.31×10^−8^, N_variants_=158). In both of the discovery and replication analyses, the association of *TREM2* was largely unaffected by the removal of low MAC variants (MAC<5) (**Supplementary Table 3**) so only moderately associated variants with MAC>4 will be highlighted as variants of interest. In the discovery dataset, there were two moderately associated variants with MAC>4, two rare missense variants (rs75932628: MAF=0.003081, *P*=2.55×10^−9^; rs143332484: MAF=0.009787, *P*=4.37×10^−5^) **(Figure 2; Supplementary Table 4)**. One of which (rs75932628) is also significant in the replication dataset (MAF=0.003278, *P*=8.71×10^−7^), whereas rs143332484 was almost moderately significant in the replication dataset (MAF=0.009947, *P*=1.30×10^−4^). The most significant variant (rs75932628) was a genome-wide significant missense variant that causes an amino acid substitution from arginine to histidine (p.R47H (TREM2-201)) in exon 2 of *TREM2*. This variant had a CADD score of 26.1, a REVEL score of 0.335 and was identified as the most strongly associated variant in *TREM2* in Sims *et al*. (2017)^17^. rs143332484 has been associated with reduced ligand affinity, signalling response, and phagocytosis of lipoprotein in microglia^18^. The other missense variant (rs143332484) also causes an amino acid substitution from arginine to histidine (p.R62H (TREM2-201)) in exon 2 of *TREM2*, however this variant is predicted to be less impactful (CADD=9.7, REVEL=0.039) and was not identified in the replication dataset, suggesting that it may not be a variant of interest. One additional moderately associated variant was found in the replication dataset (rs104894002: MAF=4.56×10^−5^, *P*=8.86×10^−6^), a pLOF variant which causes a stop gain (p.Q33S (TREM2-201)) in exon 2 and has been previously identified in AD cases^19^. *TREM2* is a well characterised AD gene and encodes a protein which is known to impact microglia anti-inflammatory response^18,20^.

**Figure 2:**
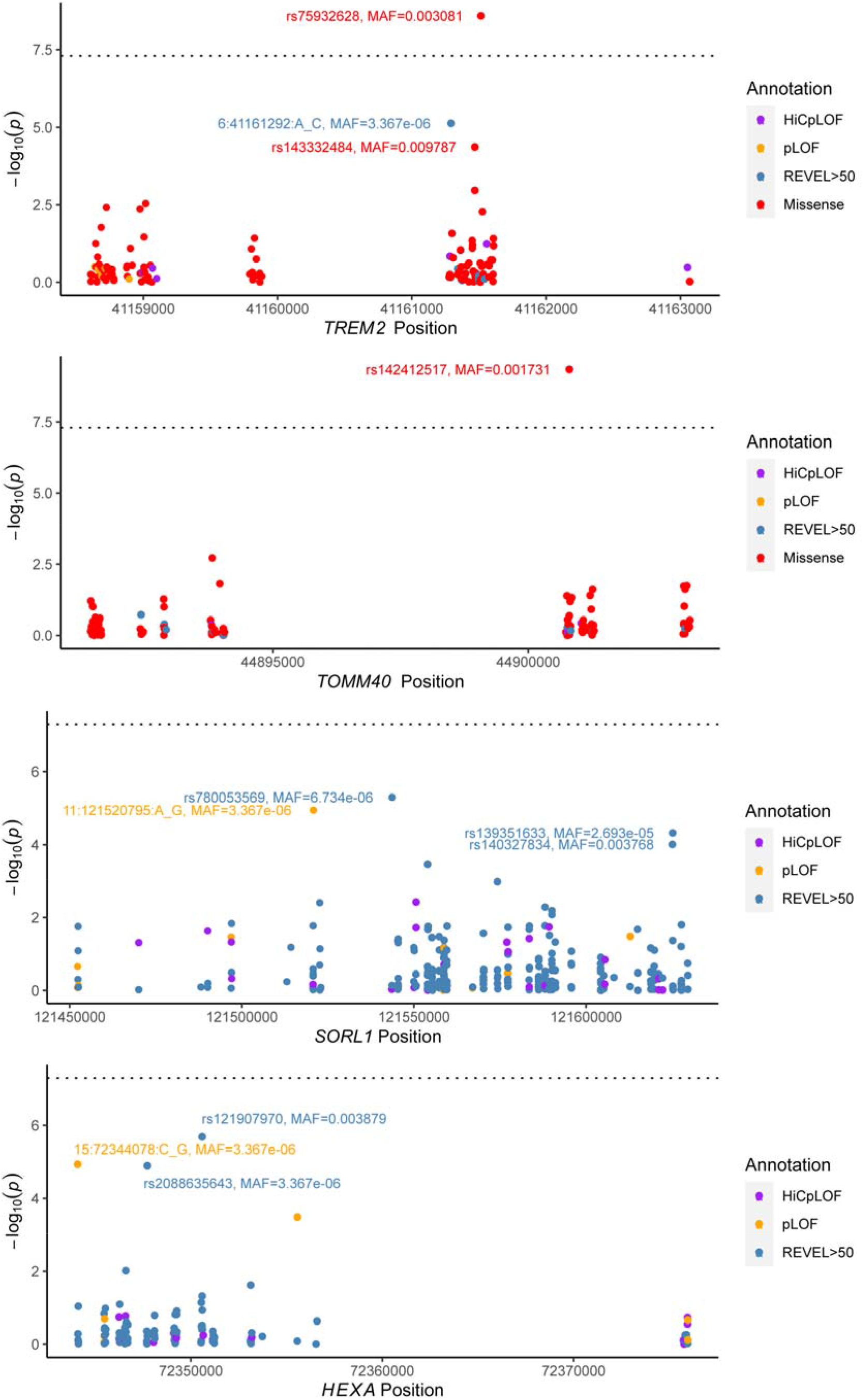
The single variant associations of variants which mapped to *TREM2, TOMM40, SORL1*, and *HEXA*. Each point represents a variant and is coloured based on functional annotation (HiCpLOF, pLOF, REVEL>50, missense). Missense variants were not plot for *SORL1* and *HEXA* because the most significant result for these genes was in the pLOF+REVEL>50 analysis.

*SORL1* was significantly associated with proxy AD/dementia when restricting to pLOF and high confidence missense variants (*P*=1.75×10^−9^, N_variants_=279). *SORL1* is a known AD gene and encodes a protein important in amyloid precursor protein processing^11^. There were two moderately associated variants with MAC>4 in *SORL1*; two high confidence missense variant (rs139351633: MAF=2.69×10^−5^, *P*=4.75×10^−5^; rs140327834: MAF=0.003768, *P*=9.81×10^−5^) (**Figure 2; Supplementary Table 4**). rs139351633 (REVEL=0.607) and rs140327834 (REVEL=0.568) affect the fibronectin type III and LDL-receptor class B regions respectively. Both of these variants were present in the replication dataset but neither were moderately associated with proxy AD/dementia (rs139351633: MAF=2.53×10^−5^, *P*=0.073; rs140327834: MAF=0.00397, *P*=0.013). Neither variant was predicted to have an impact on the protein and rs140327834 has been previously categorized as ‘likely benign’^11^. The most significant variant in the discovery dataset (rs780053569: MAF=6.73×10^−6^, *P*=4.99×10^−6^) was found in two individuals and was a missense variant (CADD=26.2; REVEL=0.592) which causes an amino acid substitution from serine to leucine (p.S602L (SORL1-201)) in a conserved region. This conserved region is the amyloid-beta binding region (vacuolar protein sorting 10 domain) of *SORL1*^11^. One additional variant was identified in the replication dataset (rs1200389078: MAF=1.01×10^−5^, *P*=1.28×10^−5^), this variant is a high confidence missense variant (p.S440L (SORL1-201)).

This variant is also in the vacuolar protein sorting 10 domain of *SORL1*^11^, but the impact of this substitution is not known. The minor alleles of all of moderately associated variants were positively associated with an increased proxy AD/dementia score which suggests that the missense or pLOF effects of the variants are associated with increased AD/dementia risk.

### Comparison to previously identified AD genes

We successfully replicated the rare variant based association of *TREM2, SORL1*, and the *APOE* locus (*TOMM40*). However, we were unable to replicate previous associations highlighted in Hoogmartens *et al*. (2021)^8^ and in Lord *et al*. (2014)^9^ with *APP, PSEN1, PSEN2, ABCA7, BIN1, UNC5C, AKAP9, NOTCH3, CLU, PLCG2, PLD3, ADAM10*, and *ABI3* (**Supplementary Table 5**). Only three of these genes have nominal significance in any of the variant aggregation analyses (pLOF+missense: *ABCA7*: *P*=1.56×10^−3^; *ABI3*: *P*=4.3×10^−3^; *PSEN1*: *P*=0.04). *ABCA7, SORL1*, and *TREM2* are the most frequently reported genes in rare-variant association studies of AD^8^, especially in recent exome-wide associations of unrelated individuals^4,5,10,21^. Two of the three most frequently identified genes in recent exome-wide association studies in unrelated individuals (*TREM2* and *SORL1*) were significantly associated with proxy AD/dementia in this study and the third gene (*ABCA7*) was nominally significant.

We aggregated the variants from these previously identified genes (excluding genes significant in this study) into a gene-set and performed SKAT-O analyses across the four variant categories using this gene-set. Three of the gene-set analyses were nominally significant (HiC pLOF: *P*=0.0096, N_variants_=292; pLOF: *P*=0.013, N_variants_=362; pLOF+REVEL: *P*=0.0015, N_variants_=1653) with the analysis using pLOF and missense variants not being nominally significant (pLOF+missense: *P*=0.16, N_variants_=5467). The P-values of all of the gene-set analyses were lower than the median P-value of the set of genes included in the gene-set (HiC pLOF=0.23, pLOF=0.45, pLOF+REVEL=0.22, pLOF+missense=0.53).

However, the gene-set P-values were approximately the same as the P-value of the most significant gene (*ABCA7*) included in that analysis, except from the pLOF+missense analysis which was considerably higher (*ABCA7*: HiC pLOF=0.02, pLOF=0.012, pLOF+REVEL=0.0017, pLOF+missense=0.0016). This suggests that the gene-set association signal in the nominally significant gene-set analyses may be mostly attributed to *ABCA7*. After removing *ABCA7* variants from the gene-set, none of the associations between the gene-set and proxy AD/dementia across the 4 variant categories are nominally significant (HiC pLOF: *P*=0.12, N_variants_=220; pLOF: *P*=0.59, N_variants_=281; pLOF+REVEL: *P*=0.14, N_variants_=1195; pLOF+missense: *P*=0.69, N_variants_=4431). We also performed gene-set analyses using gene-sets composed of genes implicated by common variants and found that *ABCA7* was the only gene driving the association between the gene-sets and proxy AD/dementia (**Supplementary Note**).

## Discussion

We performed a series of genome-wide variant aggregation analyses in 148,508 unrelated individuals of European ancestry included in the UKB to identify genes associated with proxy AD/dementia. We successfully identified 3 known AD genes (*TREM2, TOMM40*, and *SORL1*), and identified one gene not previously associated with AD (*HEXA*). All of these genes, except *HEXA*, were also significantly associated with proxy AD/dementia in a replication dataset consisting of 197,506 unrelated individuals of European ancestry included in the UKB. The role of *TREM2* and *SORL1* is well known in AD^9,11,18^ and the loss of significance of *TOMM40* after condition on APOE ε4 alleles suggests that the association of *TOMM40* is connected to the well-established *APOE* locus. *HEXA* is a known risk gene for a rare neurodegenerative disease (Tay-Sachs disease)^22^. However, neither *HEXA* nor the highlighted variant were nominally associated with proxy AD/dementia in the replication dataset, which suggests that *HEXA* is not a gene associated with AD/dementia. The initial association in the discovery dataset may have been a false positive or caused by misreporting of Tay-Sachs disease as AD/dementia in individuals included in the discovery dataset.

Across these analyses, we showed that the proxy phenotype can capture some AD gene associations identified from rare variant gene association studies of clinically diagnosed AD patients (*SORL1* and *TREM2*)^4,5,10,21^. However, we failed to identify further genes identified in previous rare variant analyses in clinically diagnosed AD patients^8^. This may be due to the differing study design between this current study and the design of the studies which initially identified these genes. The previously identified genes were found across different studies with different designs, including studies of family cohorts, unrelated individuals, exome-wide sequencing, whole gene sequencing, individual variant discovery, and variant aggregation. Previous exome-wide association studies of AD which performed similar variant aggregation as this study identified *ABCA7, SORL1*, and *TREM2*^4,10^ as genes associated with AD. We were able to replicate the significant association of *SORL1* and *TREM2*, and did find some limited support for *ABCA7*.

A limitation of the proxy phenotype is that it is a less well-defined phenotype largely based on self-reported data. The question used to define parental status does not distinguish between AD and dementia, which introduces heterogeneity in the phenotype definition. The interpretation of proxy AD/dementia associated genes not previously associated with clinically diagnosed AD should be cautious as the heterogeneity of the phenotype may highlight dementia related genes rather than AD specific genes. However, a strength of the proxy phenotype is that it allows for the inclusion of more individuals in the study by utilizing population cohorts. The inclusion of more individuals is particularly beneficial to rare variant analyses as it increases the likelihood of identifying rare variants with large impacts on protein function. The results of this study support the conclusion from previous studies^12–14,23^, which found that the proxy phenotype can capture AD genetic contributors and provide value to genetic studies of AD. We extend this to show that the proxy phenotypes can capture rare variants of interest to AD. It is important to note that variants and genes associated with proxy AD/dementia should be further validated in clinically diagnosed AD cohorts and proxy phenotypes are complementary to, and not a substitute for, well powered studies in clinically diagnosed cohorts.

## Methods

### Sample overview

This study performed exome-wide variant aggregation analyses using genetic data from 148,508 UKB participants of European ancestry. The UKB is a large population-based biobank which includes 503,325 individuals^24^. Individuals were selected for participation between 2006 and 2010. Invited individuals were between 40 and 69 years old, registered with the National Health Service, and living within 25 miles of one of the study research centres. Various data were collected from the individuals, including questionnaire answers, medical records, and genetic data. Of the 148,508 participants included in this analysis, 81,835 were female (55.1%), 66,673 were male (44.9%) and the median age was 58. We also used exome data from the remaining 197,506 unrelated individuals of European ancestry in replication analyses. In this dataset, 91,374 individuals were male (46.3%) and 106,132 were females (53.7%). The median age of the replication cohort was 58. All participants provided written informed consent; the UKB received ethical approval from the National Research Ethics Service Committee North West-Haydock (reference 11/NW/0382), and all study procedures were in accordance with the World Medical Association for medical research. Access to the UK Biobank data was obtained under application number 16406.

### Phenotype Definition

The proxy phenotype is a quantitative phenotype ranging from 0-2, where individuals with a higher score are considered to be at higher risk of developing AD/dementia based on their own diagnosis and the diagnoses of their parents. The construction of the proxy score has been described previously^14^. In brief, individuals in the UKB that report an “Alzheimer’s disease/dementia” diagnosis in either parent (data fields 20107 and 20110) are given a phenotype value based on the number of parents who have had a diagnosis. Individuals who report an AD diagnosis for themselves or have medical records reporting an AD diagnosis (ICD10 codes G30, G300, G301, G308, G309, F00, F000, F001, F002, F009 in data fields 41270, 41202 and 41204; accessed 15/04/2020) are given the same score as individuals with two parents with AD/dementia diagnoses. The contributions of parents without AD/dementia diagnoses were weighted by their age (100-age of parent/100; capped at 0.32 per parent), with older parents without AD/dementia down-weighted relative to younger ones. This resulted in 126,428 individuals with no AD diagnoses and no affected parents (proxy score <1), 20,728 individuals with one affected parent, 1075 individuals with two affected parents, and 277 individuals with an AD diagnosis themselves.

### Variant sequencing and quality control

Exome sequencing was performed for 200,643 participants of the UK Biobank study by a partnership of eight biopharmaceutical companies^25,26^. Sequencing occurred in two batches, with the first ∼50,000 individuals (UKB 50k) selected for completeness of phenotypic data and the presence of respiratory disorders of interest, and the next batch (UKB 150k) randomly selected from the larger sample of ∼500,000 individuals. Targeted regions of the exome (39Mbp in total, including 100bp flanking each gene target) were captured using the IDT xGen Exome Research Panel v1.0 with dual-indexed 75 × 75 bp paired-end reads on the Illumina NovaSeq 6000 platform using S2 (UKB 50k) and S4 (UKB 150k) flow cells.

Raw sequence reads were mapped to the GRCh38 reference genome using the OQFE protocol, followed by duplicate read marking, variant calling with DeepVariant, and filtering/merging with GLnexus. Full details of the protocol and settings are provided by Szustakowski *et al*. (2021)^25^. The resulting joint variant call file released by UKB included a total of 17,981,897 variants, with greater than 20x average coverage of 95.6% of sites in the target region. Our inspection of the data showed consistency with quality control recommendations^27^ (e.g. all samples had a transition/transversion [Ti/Tv] ratio between 2.96 and 3.21 [*M* = 3.05] for known variants; all samples had between 47,000 and 72,000 total SNPs [*M* = 54,932] in the targeted plus flanking regions). We additionally filtered the set of variants released by UKB to exclude autosomal variants with missingness > 5% (n = 343,110), variants with a minor allele count (MAC) of 0 (n = 81,027), duplicates based on position and alleles (n = 28,005), and variants outside of the targeted exome capture regions for which coverage and other quality metrics were not optimized (n = 8,800,694). After quality control, a total of 8,700,920 variants were available, of which 6,805,307 rare variants (MAF<0.01) were selected for annotation.

Array-based genotypes were also available from these same samples^28^. We used indicators of genetic kinship from these data, as provided by UKB (field id 22021) to exclude 3^rd^ degree or closer relatives. We also used these genotypes to empirically assign individuals to ancestral continental populations based on their similarity to the 1000 Genomes reference panel ancestries, and to calculate within-ancestry principal components, as described in detail by Jansen *et al*. (2019)^14^. Sample exclusion based on relatedness, ancestry, and withdrawn subjects resulted in 159,660 participants of European ancestry with exome sequencing data available for analysis. A proxy phenotype could be calculated for 148,508 individuals with available exome sequence data and these individuals were used in the analyses.

### Variant annotation with VEP

Variants were annotated with Ensembl variant effect predictor (VEP) v100.4^29^ using Ensembl version 100 data. pLOF variants were annotated using the LOFTEE plugin^30^ (github commit 2df8880). Missense variants were annotated using the REVEL v1.3 plugin^31^. Variant categories were determined based on the predicted impact of the minor allele on the gene. Four categories were created; HiC pLOF, pLOF, pLOF+REVEL>50, and pLOF+missense. HiC pLOF represents the variants deemed as high confidence loss-of-function variants by LOFTEE. pLOF represents all predicted loss-of-function variants identified by any of the following gene consequences: start_lost, stop_lost, frameshift_variant, stop_gained, splice_donor_variant, splice_acceptor_variant, or transcript_ablation. pLOF+REVEL includes all of the variants in the pLOF category plus missense variants with a REVEL score >50. This REVEL threshold was chosen because it captured 75% of disease variants and ∼11% of neutral variants in Ioannidis *et al*. (2016)^31^. The pLOF+missense category included all of the pLOF variants and all missense variants with a REVEL >=0.

### Variant aggregation analyses

Variants were aggregated in SKAT-O. (v1.3.2.1) analyses, an optimal unified test which combines burden and kernel-based tests to maximise power^15^. Only rare variants (MAF<0.01) were included in the analyses. The variants were aggregated with default weights within their mapped genes. Four SKAT-O analyses were performed, one for each of the variant categories (HiC pLOF, pLOF, pLOF+REVEL>50, and pLOF+missense). These four categories were chosen to restrict the variant aggregation to variants with likely impact on genes to aid interpretation of significantly associated genes. The analyses were performed using batch (UKB50k vs UKB150k), sex, age, and the first 10 ancestry principal components as covariates. The variant aggregation analysis was a two-sided test. Genes with low cumulative allele frequency (<0.0001) were removed to prevent genes with very few variants from biasing the analyses. The genomic inflation factors were calculated based on the P-values of each analysis. A variant burden test using only synonymous variants was performed with the same method as the other analyses in order to compare genomic inflation factors. Genes were considered significant after Bonferroni correction for the number of genes in each test and the number of variant categories (four). For the significantly associated genes, we repeated the SKAT-O analyses except with singletons and variants with MAC<5 removed to see how singletons and low MAC variants impacted the association of the significant genes.

### Replication analyses

We used the exome data of the remaining 197,506 unrelated individuals of European ancestry to perform the replication SKAT-O analyses in the four significant genes and four BH-FDR genes. The exome data was obtained in plink binary format from the final release folder provided by the UKB. The methods used to create this data have been described in Backman *et al*. (2021)^32^. Ancestry assignment and relatedness was calculated as described above. We removed variants where less than 90% of all genotypes for that variant had a read depth less than 10 (ukb23158_500k_OQFE.90pct10dp_qc_variants.txt). Variants with MAF<0.01 were removed. Then, the variant annotation and variant aggregation analyses were performed as described above. Batch was not included as a covariate as no individuals from the UKB50K or UKB150k datasets were included in the replication dataset. The replication dataset consisted of 171,526 individuals with no AD diagnoses and no affected parents (proxy score <1), 24,485 individuals with one affected parent, 1086 individuals with two affected parents, and 409 individuals with an AD diagnosis themselves.

### Investigating single variants

In order to highlight variants of interest, PLINK (v1.9b_6.17)^33,34^ linear regression was performed to identify the association of the variants within significant genes identified in the SKAT-O analyses (*TREM2, TOMM40, SORL1* and *HEXA*) with the proxy phenotype. The variant association analysis was limited to the variants included in the SKAT-O analyses. The linear regression was performed using batch (in discovery dataset), sex, age, and the first 10 principal components as covariates. The impact of the variants on the proteins was predicted using mutfunc^35^. CADD^36^ score and amino acid substitutions were identified during the initial variant annotation by VEP. For each of the amino acid substitutions reported in the main text, the Ensembl transcript name of one transcript affected by the amino acid substitution is included in brackets. All of the corresponding Ensembl transcript names and IDs of the amino acid substitutions reported in the text or **Supplementary Note** are included in **Supplementary Table 6**. All plots were generated in R using ggplot2^37^ or base R^38^ functions. The four genes that showed significance in the variant aggregation analyses were tested again in the same variant aggregation analyses except with singletons (variants present in only one individual) excluded, variants with low MAC (<5) excluded, and then with moderately associated variants (*P*<1×10^−4^) excluded.

### Variant level quality metrics

Quality metrics (mean sequencing depth and variance, missing rate, and allele quality) of the moderately associated variants in the significant genes are available in **Supplementary Table 4**. Among the moderately associated variants, the mean read depth ranged from ∼17-27, the missing rate was less than 1×10^−4^, and the allele quality (phred scale) ranged from 36-58 (approximately 99.9-99.999% base call accuracy). Quality metrics for all variants which were included in the analysis where *HEXA* was significantly associated with the proxy phenotype are available in **Supplementary Table 7**. Among the *HEXA* variants (N=121) the mean depth ranged from ∼15-36, the missing rate was less than 1×10^−3^, and the allele quality (phred scale) ranged from 36-58.

### Comparison to previously identified AD genes

We looked at the association signal in genes previously associated with AD through rare variants and common variants. Rare variant genes were chosen based on their presence as replicated genes with rare variants reviewed in Hoogmartens *et al*. (2021)^8^ and their presence as genes with rare variants reviewed in Lord *et al*. (2014)^9^. The common variant genes were selected from Table 1 (‘known loci’) and Table 2 (‘new loci’) from the most recent AD GWAS, Bellenguez *et al*. (2022)^39^. We selected genes from the ‘Known locus’ column of Table 1 and the ‘Gene’ column from Table 2. If multiple genes were highlighted in a locus name, all of those genes were included in the gene-set. We aggregated all of the variants in each variant category which map to the genes present in the rare variant genes to make 4 gene-sets. We then tested that gene-set using SKAT-O, as previously described, where all the variants in the gene-set were aggregated together. The same covariates were used as in the gene analyses. We repeated this gene-set analysis except with gene-sets defined by the common variant genes in ‘known loci’ and ‘new loci’. Due to memory limitation, we had to perform the gene-set analysis for the genes from the ‘known loci’ separately from the ‘new loci’.

### Data availability statement

The gene-level summary statistics for all four models are available at https://github.com/dwightman/UKBrarevariant and will be made available at https://ctg.cncr.nl/software/summary_statistics after publication. The individual level exome and phenotype data are available through the UK Biobank to approved researchers.

Researchers can apply to access the UK Biobank data through https://www.ukbiobank.ac.uk/enable-your-research.

### Code availability statement

The code used in this study is available at https://github.com/dwightman/UKBrarevariant.

## Data Availability

The gene-level summary statistics for all four models are available at https://github.com/dwightman/UKBrarevariant and will be made available at https://ctg.cncr.nl/software/summary_statistics after publication. The individual level exome and phenotype data are available through the UK Biobank to approved researchers. Researchers can apply to access the UK Biobank data through https://www.ukbiobank.ac.uk/enable-your-research.

## Acknowledgements

DP was funded by The Netherlands Organization for Scientific Research (NWO VICI 453-14-005), NWO Gravitation: BRAINSCAPES: A Roadmap from Neurogenetics to Neurobiology (Grant No. 024.004.012), and a European Research Council advanced grant (Grant No, ERC-2018-AdG GWAS2FUNC 834057). DW was funded by NWO Gravitation: BRAINSCAPES: A Roadmap from Neurogenetics to Neurobiology (Grant No. 024.004.012). IEJ was funded by NWO Gravitation: BRAINSCAPES: A Roadmap from Neurogenetics to Neurobiology (Grant No. 024.004.012). JES was supported by funding from the Amsterdam Neuroscience Alliance Project. CdL was funded by F. Hoffmann-La Roche AG. The research has been conducted using the UK Biobank Resource (application no. 16406). Analyses were carried out on the Genetic Cluster Computer hosted by the Dutch National computing and Networking Services SurfSARA.

## Author contributions

DPW and JES analysed the data. DPW and JES wrote the manuscript. DPW, IEJ, CAdL, and DP designed the analysis plan. IEJ, CAdL, and DP supervised the project.

