## Supplementary Note for "Rare Variant Aggregation in 148,508 Exomes Identifies Genes Associated with Proxy Alzheimer’s disease/Dementia"

Supplementary Results

Subthreshold genes

In order to investigate if there are any genes with strong associations with proxy AD/dementia signal which do not pass the Bonferroni correction threshold in the discovery dataset, we selected genes at Benjamini–Hochberg false-discovery rate (BH-FDR) of 10% in each of the 4 variant category SKAT-O analyses (**Supplementary Table 8**). No genes were identified in either of the LOF categories, suggesting that the signal in these analyses was very sparse. Three additional genes were identified in the pLOF+REVEL>50 analysis; *FBXW4* (P=6.47x10^-6^, N_variants_=38), *BPIFB3* (P=1.80x10^-5^, N_variants_=13), *HS3ST2* (P=2.39x10^-5^, N_variants_=37). Removing singletons from these analyses does not considerably affect the results (*FBXW4*: P=3.93x10^-6^, N_variants_=20; *BPIFB3*: P=5.92x10^-6^, N_variants_=8; *HS3ST2*: P=2.25x10^-5^, N_variants_=11); however, removing all variants with MAC<5 does affect *BPIFB3* and *HS3ST2* considerably (*FBXW4*: P=4.93x10^-6^, N_variants_=10; *BPIFB3*: P=0.57, N_variants_=3; *HS3ST2*: P=0.65, N_variants_=1).

*FBXW4* encodes F-box/WD repeat-containing protein 4, a member of a family of proteins that contain F-box domains to recruit ubiquitin ligase and WD domains to recruit proteins to be labelled with ubiquitin for degradation^1^. *FBXW4* has been connected to acute myeloid leukemia^1^ but further disease associations are not well documented. Two other members of the F-box/WD family have been connected to AD through interaction with presenilin 1 (FBXW7)^2^ and a mouse model (*FBXW11*)^3^; however, the connection between *FBXW4* and AD is not well characterized. *BPIFB3* encodes BPI fold containing family B member 3, the function of this protein is not well characterized but it appears to impact infection response^4,5^. As with *FBXW4*, the connection between *BPIFB3* and AD is not clear. *HS3ST2* encodes heparan sulfate-glucosamine 3-sulfotransferase 2, an enzyme which adds 3-O-sulphated domains to heparin sulfates^6^. These 3-O-suphated domains have been hypothesized to contribute to AD by influencing the abnormal phosphorylation of tau^6^. Glycoproteins with heparan sulfates attached (Heparan sulfate proteoglycans) have been previously linked to many pathways that contribute to AD^7,8^. There appears to be some evidence for the role of *HS3ST2* in AD but further replication of the experimental work connecting *HS3ST2* to AD is needed. *FBXW4*, *BPIFB3*, and *HS3ST2* were not nominally significant in the replication dataset (**Supplementary Table 8**), which suggests that there is little evidence from rare variants to support these genes as AD/dementia genes.

Four additional genes were identified in the pLOF+missense analysis; *SORL1* (P=2.58x10^-6^; N_variants_=782), *HEXA* (P=3.82x10^-6^; N_variants_=196), *BFAR* (P=3.84x10^-6^; N_variants_=154), and *PVR* (P=2.31x10^-5^; N_variants_=158). *SORL1* and *HEXA* passed the Bonferroni correction threshold in the pLOF+REVEL>50 analysis and are discussed elsewhere. *PVR* is a gene in the *APOE* region and after conditioning on the *APOE* ε4 allele, *PVR* was not even nominally significant (P=0.36). This leaves *BFAR* as one interesting gene not previously highlighted in other analyses. *BFAR* was largely unaffected by the removal of singletons (*BFAR*: P=4.40x10^-6^, N_variants_=68) and variants with MAC<5 (*BFAR*: P=2.90x10^-6^, N_variants_=38). *BFAR* encodes for bifunctional apoptosis regulator (BAR), this protein sensitizes neurons to cell death if down regulated^9^. BAR binds to Bcl-2^10^, a protein which inhibits apoptosis^11^ and has been implicated in AD^12^. It is possible that missense mutations in *BFAR* could limit the anti-apoptosis ability of BAR through reduced interaction with Bcl-2. However, *BFAR* was not nominally significant in the replication dataset (**Supplementary Table 8**), which suggests that there is little evidence from rare variants to support *BFAR* as an AD/dementia gene.

We then used all of the selected genes at BH-FDR rate of 10% in each of the 4 analyses to perform gene-set enrichment using FUMA GENE2FUNC^13^. This analysis identified 4 gene-sets significantly enriched with the selected genes after Bonferroni correction for the number of tested gene-sets. One MSigDB gene-set and three gene-sets based on GWAS catalog reported genes. The MSigSB gene-set based on genes which bind to low-density lipoprotein was significantly enriched (GOMF_LOW_DENSITY_LIPOPROTEIN_PARTICLE_BINDING: P_Bonferroni_= 0.045); however, only *SORL1* and *TREM2* overlapped with this gene-set. The GWAS catalog gene-sets were based on reported gene associations with AD phenotypes (Alzheimer's disease or family history of Alzheimer's disease: P_Bonferroni_= 6.43x10^-9^; Alzheimer's disease: P_Bonferroni_= 2.90x10^-9^; Alzheimer's disease (late onset): P_Bonferroni_= 8.45x10^-6^). These results were driven by *TREM2*, *PVR*, *TOMM40*, and *SORL1*.

Gene-set Analyses

In order to identify biological processes implicated by rare variants associated with proxy dementia, we performed gene-set analyses on 9769 gene-sets from MSigDB v7.0. Due to memory constraints we could only analyse gene-sets with <10,000 variants. We used the same SKAT-O model as with the gene analysis except all variants which mapped to the genes in the gene-set were aggregated together. After Bonferroni correction for 9769 gene-sets, we identified 33 significant gene-sets (**Supplementary Table 9**). All of these associations were robust to the exclusion of all variants in the larger *APOE* region (GRCh38: 19:40000000-50000000). However, none of these associations were robust to the exclusion of the variants in the larger *APOE* region (which includes *TOMM40*) and variants present in the 4 significant genes (*TREM2*, *HEXA*, *SORL1*, *TOMM40*). After removal of these variants only two of the significant gene-sets had even nominal significance. These two gene-sets (GO_bp:go_leukocyte_activation_involved_in_inflammatory_response and GO_bp:go_astrocyte_activation) each contained three genes (outside of the *APOE* region genes and *TREM2*, *HEXA*, *SORL1*, *TOMM40*) with nominal significance (P<0.05) in the pLOF+missense analyses. The three nominally significant genes in GO_bp:go_leukocyte_activation_involved_in_inflammatory_response (*P*=0.034, N_variants_=7680) were *PTPRC* (*P*=0.0015, N_variants_=387), *MAPT* (*P*=0.018, N_variants_=275), and *GRN* (*P*=0.024, N_variants_=244). The three nominally significant genes in GO_bp:go_astrocyte_activation (*P*=0.033, N_variants_=4722) were *MAPT* (*P*=0.018, N_variants_=275), *GRN* (*P*=0.024, N_variants_=244), *PSEN1* (*P*=0.040, N_variants_=113). This suggests that the associated gene-sets were almost entirely driven by variants in *TREM2*, *HEXA*, and *SORL1*, and that the gene-set analysis was not capturing additional information not found from the gene level analysis.

*TOMM40* Variants

*TOMM40* was significantly associated with proxy AD/dementia when all missense and pLOF variants were included in the variant aggregation analysis (*P*=2.39x10^-10^, N_variants_=119). There was one variant of interest in TOMM40; a significant rare missense variant (rs142412517; MAF=0.001731; *P*=4.43x10^-10^) **(Figure 2; Supplementary Table 4)**. This variant was also the only moderately associated variant in the replication dataset (*P*=5.05x10^-12^, MAF=0.00181). This missense variant (CADD=25.4; REVEL=0.238) causes an amino acid change from arginine to tryptophan (p.R239W (TOMM40-201)) in a conserved region, but the impact on the protein is unclear. This variant is in very low LD (<0.01) with the APOE ε4 variants (rs7412-C, rs429358-C), but when conditioning for APOE ε4 status based on these two variants, *TOMM40* was no longer significantly associated with the phenotype (*P*=0.043) **(Supplementary Table 2)**. No other significant gene lost significance when conditioned on APOE ε4 status. The low number of variants in the HiC LOF, pLOF, and pLOF+REVEL>50 analyses caused this gene to fail the cumulative frequency threshold (0.0001) so *TOMM40* was not included in these analyses. The influence of missense variants and *APOE* ε4 variants on the association of *TOMM40* suggests that the impact of the variant on *TOMM40* protein structure is not the primary driver of the association.

*HEXA* Variants

*HEXA* was significantly associated with the phenotype when restricting to pLOF and high confidence missense variants (*P*= 2.13x10^-7^, N_variants_=121). There was one moderately associated variant with a MAC>1 in *HEXA*, a high-confidence missense variant (rs121907970: MAF=0.003879, *P*=2.07x10^-6^) (**Figure 2; Supplementary Table 4**). This variant is a missense variant (CADD=25.1; REVEL=0.925) which causes an amino acid substitution from arginine to tryptophan (p.R247W (HEXA-209)/p.R258W (HEXA-206)). This amino acid change has been linked to a reduced activity of Hex A, however this reduction in activity is only in regards to an artificial substrate used to measure Hex A activity and does not affect in situ activity^14^. In the discovery dataset, at least one risk allele of rs121907970 was present in 6 of the 277 (2.17%) individuals with an AD diagnosis, 14 of the 1075 (1.30%) individuals with two parents with an AD/dementia diagnosis, 195 of the 20,728 (0.94%) individuals with one parent with an AD/dementia diagnosis, and 933 of the 126,428 (0.74%) individuals with no AD diagnosis and no parent with an AD/dementia diagnosis. By ordering the outcome from least to most AD/dementia risk (no diagnosis and no affected parent → 1 affected parent → 2 affected parents → AD diagnosis), the individuals with higher AD/dementia risk were more likely to have at least one copy of the rs121907970 risk allele (two-sided ordinal chi-square/linear-by-linear association test: Z=4.278, P=1.89x10^-5^). However, rs121907970 was not even nominally significant in the replication dataset (*P*=0.46, MAF=0.0038) and the risk allele of this variant was not more likely to be present in individuals with higher AD/dementia risk (two-sided ordinal chi-square/linear-by-linear association test: Z=1.0089, P=0.313). This suggests that this variant is unlikely to be truly associated with AD/dementia. *HEXA*, along with *HEXB*, encodes the protein β-hexosaminidase (Hex) which degrades GM2 gangliosides^15^. The accumulation of GM2 gangliosides leads to 3 rare neurodegenerative lysosomal storage disorders (Tay-Sachs, Sandhoff disease, and GM2 activator deficiency). GM2 accumulation is associated with neuronal structural change, microglial activation and response, and neuroglial dysfunction. There is also some evidence to support the colocalization of Hex with amyloid-beta plaques in the brains of AD patients^16^. However, given that *HEXA* and the sole highlighted variant were not even nominally significant in the replicated cohort, it is unlikely that HEXA is truly associated with proxy AD/dementia.

Comparison to previously identified AD genes through common variants

We further looked at whether genes identified in common variant GWAS analyses were enriched in association signal. We selected genes highlighted in the ‘Known locus’ column of Table 1 and the ‘Gene’ column from Table 2 of Bellenguez *et al*. (2022)^17^ to act as AD genes previously associated through common variants. After excluding the genes implicated by the SKAT-O analyses (*APOE*, *TOMM40*, *SORL1*, *HEXA*, and *TREM2*) and the HLA region, 14 of the 73 AD genes had nominal significance (P<0.05) in at least one of the four SKAT-O analyses (**Supplementary Table 10**), including *ABI3* and *ABCA7* which were also implicated as AD genes through previous rare variant analyses. However, none of these genes reach significance after Bonferroni correction for the total number of common variant genes. To see if the association signal in these genes would combine in aggregate, we performed a gene-set analysis similar to the rare variant genes gene-set analysis described in the **Results** section of the main text, except using the genes identified through common variant GWAS. Due to memory limitations, we were unable to combine the variants in all of the common variant genes so we created two gene-sets, one for the genes in Table 1 (known loci) and one for the genes in Table 2 (new loci). The new loci gene-set was not nominally significant using any of the four variant categories (**Supplementary Table 10**); however, the known loci gene-set was nominally significant in the pLOF, pLOF+REVEL, and pLOF+missense variant category analyses (**Supplementary Table 10**). *ABCA7* was included in the known loci gene-set, so we tested whether these nominally significant results were robust to the exclusion of *ABCA7* variants and found that only the pLOF+missense category was nominally significant (P=0.0497) after removal of *ABCA7* variants. These results along with the results from the genes previously implicated by rare variants suggests that there is little association signal in previously implicated AD genes outside of *APOE* (*TOMM40*), *SORL1*, *TREM2*, and *ABCA7*.

Supplementary Methods

Subthreshold genes

We identified genes at Benjamini–Hochberg false-discovery rate (BH-FDR) of 10% in each of the 4 variant category SKAT-O analyses. One of these genes (*PVR*) was in the *APOE* region, so we repeated the SKAT-O analysis of this gene and included *APOE* ε4 status (based on rs7412-C and rs429358-C) as a covariate. We repeated the SKAT-O analyses for the BH-FDR genes without singletons and low MAC (<5) variants. Then using the full set of BH-FDR genes (N=9), including *TOMM40*, *HEXA*, *TREM2*, and *SORL1*, we performed gene-set enrichment using FUMA GENE2FUNC^13^ (v1.3.8). We compared the BH-FDR genes to the other genes tested in the analyses (N=18425); however, FUMA only recognized the Ensembl ID for 17,991 genes. The number of tests were adjusted using Bonferroni correction and the Bonferroni adjusted P-value cutoff was 0.05. Only gene-sets containing at least 2 overlapping genes were tested.

SKAT-O Gene-set Analyses

We performed gene-set analyses to look if sub-threshold association signal would aggregate across genes to highlight biological processes. All pLOF and missense variants were used in the gene-set analysis because this was the variant category with the highest inflation and potentially the most association signal to be aggregated. We analysed 9769 gene-sets from MSigDB v7.0, we were only able to analyse gene-sets with <10,000 variants due to memory constraints. The gene-set analysis was performed using SKAT-O in the same way as the gene analyses, except all variants which mapped to the genes in the MSigDB gene-sets were aggregated together. We then repeated the analyses of the Bonferroni corrected significant gene-sets with variants in the larger *APOE* region (GRCh38: 19:40000000-50000000) removed. We again repeated the analyses with the variants in the larger *APOE* region and the variants in the significantly associated genes from the gene analysis (*TREM2*, *HEXA*, *SORL1*, *TOMM40*) removed.
